# The evaluation of a novel digital immunochromatographic assay with silver amplification to detect SARS-CoV-2

**DOI:** 10.1101/2021.05.06.21256738

**Authors:** Yoko Kurihara, Yoshihiko Kiyasu, Yusaku Akashi, Yuto Takeuchi, Kenji Narahara, Sunao Mori, Tomonori Takeshige, Shigeyuki Notake, Atsuo Ueda, Koji Nakamura, Hiroichi Ishikawa, Hiromichi Suzuki

## Abstract

**Introduction:** Rapid antigen tests are convenient for diagnosing severe acute respiratory syndrome coronavirus 2 (SARS-CoV-2); however, they have lower sensitivities than nucleic acid amplification tests. In this study, we evaluated the diagnostic performance of Quick Chaser^®^ Auto SARS-CoV-2, a novel digital immunochromatographic assay that is expected to have higher sensitivity than conventional antigen tests.

**Methods:** A prospective observational study was conducted between February 8 and March 24, 2021. We simultaneously obtained two nasopharyngeal samples, one for evaluation with the QuickChaser^®^ Auto SARS-CoV-2 antigen test and the other for assessment with reverse transcription PCR (RT-PCR), considered the gold-standard reference test. The limit of detection (LOD) of the new antigen test was compared with those of four other commercially available rapid antigen tests.

**Results:** A total of 1401 samples were analyzed. SARS-CoV-2 was detected by reference RT-PCR in 83 (5.9%) samples, of which 36 (43.4%) were collected from symptomatic patients. The sensitivity, specificity, positive predictive value, and negative predictive value were 74.7% (95% confidence interval (CI): 64.0–83.6%), 99.8% (95% CI: 99.5–100%), 96.9% (95% CI: 89.2–99.6%), and 98.4% (95% CI: 97.6–99.0%), respectively. When limited to samples with a cycle threshold (Ct) <30 or those from symptomatic patients, the sensitivity increased to 98.3% and 88.9%, respectively. The QuickChaser^®^ Auto SARS-CoV-2 detected 34–120 copies/test, which indicated greater sensitivity than the other rapid antigen tests.

**Conclusions:** QuickChaser^®^ Auto SARS-CoV-2 showed sufficient sensitivity and specificity in clinical samples of symptomatic patients. The sensitivity was comparable to RT-PCR in samples with Ct<30.

## Introduction

The severe acute respiratory syndrome coronavirus 2 (SARS-CoV-2) has caused a global pandemic and continues to place an immense burden on healthcare systems [1] despite the introduction of effective vaccines [2]. Since rapid and accurate testing is a critical element in containing viral transmission [3], the development of reliable point-of-care testing is necessary.

Nucleic acid amplification tests (NAATs) are the gold standard for diagnosing coronavirus disease 2019 (COVID-19) because of their high diagnostic performance [4]. However, several limitations have reduced their test capacity and clinical utility, including long processing times and the need for expensive equipment and skilled staff [3]. By contrast, antigen tests are convenient and have moderate sensitivities and high specificities [4]. These tests have made it possible to diagnose COVID-19 in low-resource settings [5], despite the possibility of missing a certain proportion of infected patients [6]. Therefore, increasing sensitivity should enhance the clinical utility of antigen tests.

Quick Chaser^®^ Auto SARS-CoV-2 (Mizuho Medy, Saga, Japan) is a new antigen test based on the silver amplification method. This test uses the same reagent as FUJI DRY-CHEM IMMUNO AG Cartridge COVID-19 Ag (Fujifilm, Tokyo, Japan), and is tailored for digital immuno-chromatographic assays. The test provides results in 15 minutes when used with the QuickChaser Immuno Reader II dedicated reader (Mizuho Medy). Both the silver amplification method and digital immuno-chromatographic assays were reported to increase the sensitivity of antigen tests for the influenza virus [7]. Although Quick Chaser^®^ Auto SARS-CoV-2 is expected to have higher sensitivity than conventional antigen tests, its diagnostic performance for detecting SARS-CoV-2 has not been evaluated in clinical samples.

In this study, we evaluated the diagnostic performance of Quick Chaser^®^ Auto SARS-CoV-2 and QuickChaser Immuno Reader II with nasopharyngeal specimens, and performed comparisons with the reverse transcription PCR (RT-PCR) method.

## Methods

This study was carried out as an extension of our previous research [8] and followed a similar protocol. The investigation was performed between February 8 and March 24, 2021, at Tsukuba Medical Center Hospital (TMCH), a tertiary hospital in Ibaraki Prefecture, Japan. Nasopharyngeal samples and clinical information were gathered from individuals who had possibly contracted SARS-CoV-2. The enrolled patients were referred from 67 nearby clinics and a local public health center, and by healthcare workers at TMCH. All patients provided informed consent to participate in the study, which was approved by the ethics committee of TMCH (approval number: 2020-071).

### Procedures for sample collection and antigen test

Two nasopharyngeal samples were obtained from each patient for further testing: one with a sponge swab™ (NIPRO, Osaka, Japan) for antigen testing, and the other with FLOQSwab™ (Copan Italia S.p.A., Brescia, Italy) for the RT-PCR assay. After sample collection, antigen testing was performed immediately using the QuickChaser^®^ Auto SARS-CoV-2 and QuickChaser Immuno Reader II. FLOQSwab samples were diluted in 3 mL of Universal Transport Medium™ (UTM™) (Copan Italia) for in-house RT-PCR and reference RT-PCR.

### RT-PCR assay for SARS-CoV-2

For in-house RT-PCR, magLEAD 6gC (Precision System Science, Chiba, Japan) was used to extract and purify RNA from UTM™ samples. The samples were then transferred to Mizuho Medy for reference real-time RT-PCR. The N2 primer/probe set (Nihon Gene Research Laboratories, Miyagi, Japan) was employed for reference RT-PCR as suggested by the “Manual for the Detection of Pathogen 2019-nCoV Ver. 2.9.1” issued by the National Institute of Infectious Diseases of Japan [9]. The RT-PCR assays were performed on a Thermal Cycler Dice III (Takara Bio, Kusatsu, Japan) using One Step PrimeScript™ III RT-qPCR Mix (Takara Bio) with the following cycling conditions: reverse transcription at 52 °C for 5 min and at 95 °C for 10 s, and 45 cycles at 95 °C for 5 s and at 60 °C for 30s. The absolute viral copy number was determined by serially diluted RNA control targeting the N2 gene of SARS-CoV-2 (Nihon Gene Research Laboratories). If the in-house and reference RT-PCR showed conflicting results, GeneXpert^®^ for SARS-CoV-2 (Cepheid, Sunnyvale, CA, USA) was used to re-examine the sample for the final decision.

### Limits of detection of QuickChaser^®^ Auto SARS-CoV-2 and four commercially available rapid antigen tests

We compared the limit of detection (LOD) of QuickChaser^®^ Auto SARS-CoV-2 with those of four commercially available rapid antigen tests (Espline^®^ SARS-CoV-2, Fujirebio, Tokyo, Japan; QuickNavi™-COVID19 Ag, Denka, Tokyo Japan; Panbio™ COVID-19 Ag Rapid Test Device, Abbott Diagnostics, Illinois, USA; SARS-CoV-2 Rapid Antigen Test, Roche Diagnostics, Rotkreuz, Switzerland). Two SARS-CoV-2–positive, cryopreserved, nasopharyngeal swab specimens were serially diluted two-fold with UTM™. The diluted solution was collected with the swabs included in each antigen test kit and was added to the extraction reagent solution of each antigen test. After that, these extracted samples were dropped into test cartridges. The LOD of each antigen test was evaluated according to the measurement method described in the package insert, and results were agreed upon by three researchers.

The numbers of viral copies contained in the UTM™ samples were determined by RT-PCR with viral RNA extraction performed using a QIAamp Viral RNA Mini Kit (QIAGEN N.V., Hilden, Germany).

### Statistical analyses

The sensitivity, specificity, positive predictive value (PPV), and negative predictive value (NPV) of QuickChaser^®^ Auto SARS-CoV-2 were calculated using the Clopper and Pearson method, with 95% confidence intervals (CIs). All calculations were conducted using the R 4.0.3 software program (www.r-project.org).

## Results

During the study period, 1,416 nasopharyngeal samples were initially included. Samples with missing clinical data (n = 14) or measurement errors (n = 1) were excluded. A total of 1,401 samples were eventually analyzed.

Reference real-time RT-PCR detected SARS-CoV-2 in 83 (5.9%) of the 1401 samples. The results of reference and in-house RT-PCR were consistent in all but one sample, which was negative by in-house RT-PCR and positive by reference RT-PCR. This sample was re-evaluated by in-house RT-PCR and GeneXpert^®^ using preserved UTM. Both tests showed positive results, and the sample was finally considered to be positive for SARS-CoV-2. Of the 83 samples, 36 (43.4%) were collected from symptomatic patients, and 47 (56.6%) were obtained from asymptomatic participants. The relationship between the interval from symptom onset and the sensitivity of QuickChaser^®^ Auto SARS-CoV-2 is shown in Figure 1.

**Figure 1.**
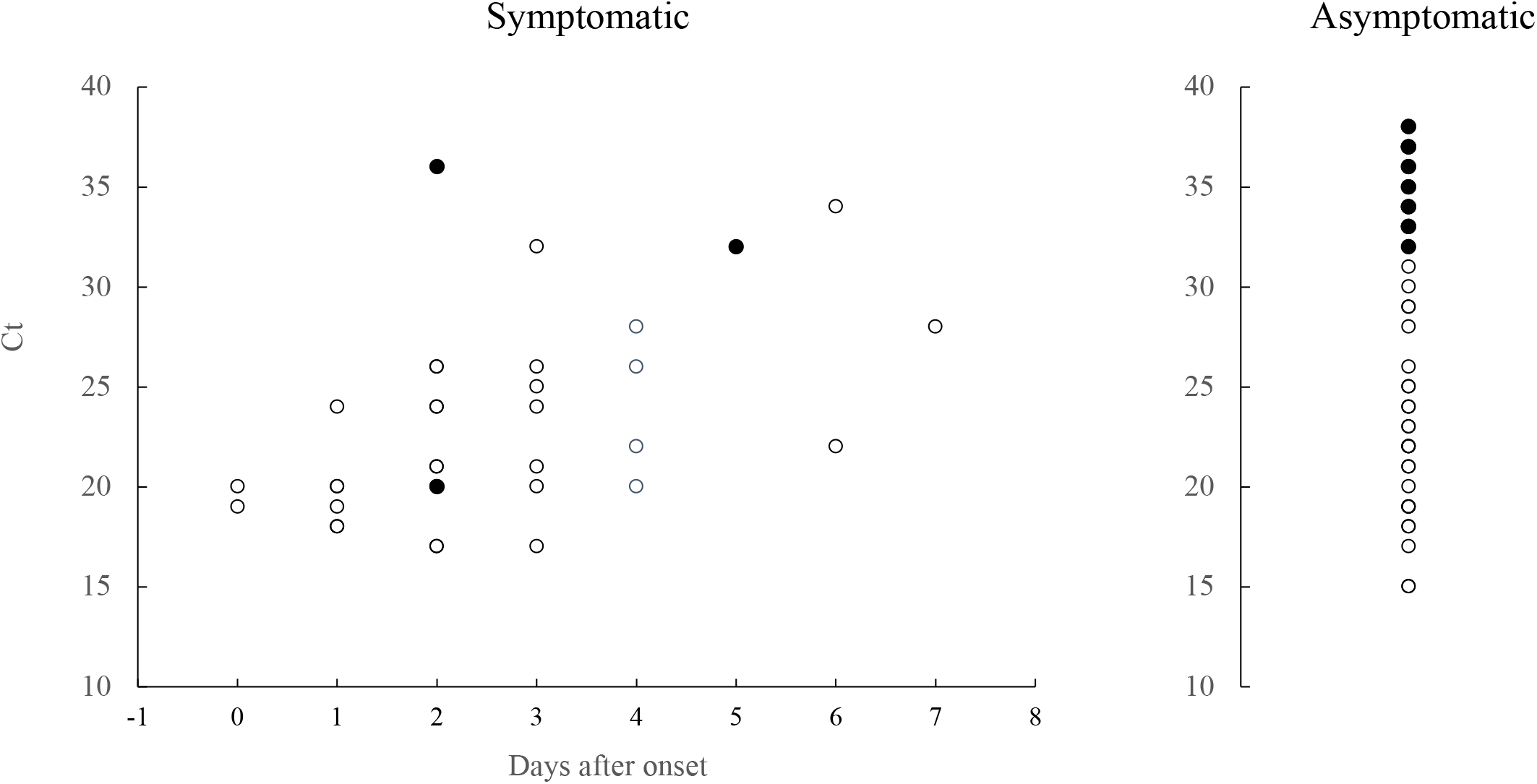
Difference in sensitivity of QuickChaser^®^ Auto SARS-CoV-2 stratified by the day after symptoms onset. Two patients with unknown onset dates were excluded. White circles indicate positive samples, and black circles indicate negative samples for the antigen test.

### Comparison of LODs

The results of LOD tests using clinical specimens are summarized in Table 1. Among the five antigen tests, the QuickChaser^®^ Auto SARS-CoV-2 had the lowest LOD: QuickChaser^®^ Auto SARS-CoV-2, 34–120 copies/test; Espline SARS^®^-CoV-2, 481–549 copies/test; QuickNavi™-COVID19 Ag, 4,394 copies/test; Panbio™ COVID-19 Ag Rapid Test Device, 1,098–1,924 copies/test; and SARS-CoV-2 Rapid Antigen Test, 549–1,924 copies/test.

**Table 1.** Limits of detection tests using nasopharyngeal swab samples.

|  | Dilution factor | Copies/test | Quick Chaser | Espline | Quick Navi | Panbio | Rapid Antigen Test |  |
| --- | --- | --- | --- | --- | --- | --- | --- | --- |
|  |  |  | 15 min | 30 min | 15 min | 15 min | 15 min | 30 min |
| Sample 1 | 20 | 8,787 | + | + | + | + | + | + |
|  | 40 | 4,394 | + | + | + | + | + | + |
|  | 80 | 2,197 | + | + | – | + | + | + |
|  | 160 | 1,098 | + | + | – | + | + | + |
|  | 320 | 549 | + | + | – | – | + | – |
|  | 640 | 275 | + | – | – | – | – | – |
|  | 1,280 | 137 | + | – | – | – | – | – |
|  | 2,560 | 69 | + | n.t. | n.t. | n.t. | n.t. | n.t. |
|  | 5,120 | 34 | + | n.t. | n.t. | n.t. | n.t. | n.t. |
|  | 10,240 | 17 | – | n.t. | n.t. | n.t. | n.t. | n.t. |
| Sample 2 | 20 | 3,849 | + | + | – | + | + | + |
|  | 40 | 1,924 | + | + | – | + | + | + |
|  | 80 | 962 | + | + | – | – | – | – |
|  | 160 | 481 | + | + | – | – | – | – |
|  | 320 | 241 | + | – | – | – | – | – |
|  | 640 | 120 | + | n.t. | n.t. | n.t. | n.t. | n.t. |
|  | 1,280 | 60 | – | n.t. | n.t. | n.t. | n.t. | n.t. |
|  | 2,560 | 30 | – | n.t. | n.t. | n.t. | n.t. | n.t. |
|  | 5,120 | 15 | – | n.t. | n.t. | n.t. | n.t. | n.t. |

+, Positive; –, negative; n.t., not tested;
QuickChaser™, QuickChaser® Auto SARS-CoV-2; Espline™, Espline® SARS-CoV-2 (Fujirebio, Tokyo, Japan); QuickNavi™, QuickNavi™-COVID19 Ag (Denka, Tokyo, Japan); Panbio™, Panbio™ COVID-19 Ag Rapid Test Device (Abbott Diagnostics Medical, Illinois, USA); Rapid Antigen Test, SARS-CoV-2 Rapid Antigen Test (Roche Diagnostics, Rotkreuz, Switzerland)

### Sensitivity, specificity, PPV, and NPV of QuickChaser^®^ Auto SARS-CoV-2

The clinical performance of QuickChaser^®^ Auto SARS-CoV-2 is summarized in Tables 2 and 3. Sixty-two of the 83 samples that were positive by reference RT-PCR were also positive by the antigen test. The sensitivity, specificity, PPV, and NPV were 74.7% (95% CI: 64.0–83.6%), 99.8% (95% CI: 99.5–100%), 96.9% (95% CI: 89.2–99.6%), and 98.4% (95% CI: 97.6–99.0%), respectively (Table 2).

**Table 2.** **Sensitivity and specificity of QuickChaser**^**®**^ **Auto SARS-CoV-2 among overall subjects**

| Overall subjects |  | Real-time RT-PCR |  |
| --- | --- | --- | --- |
|  |  | Positive | Negative |
| QuickChaser<sup>®</sup> Auto SARS-CoV-2 | Positive | 62 | 2 |
|  | Negative | 21 | 1316 |
| Sensitivity (%) |  | 74.7 (64.0–83.6) |  |
| Specificity (%) |  | 99.8 (99.5–100) |  |
| Positive predictive value (%) |  | 96.9 (89.2–99.6) |  |
| Negative predictive value (%) |  | 98.4 (97.6–99.0) |  |

RT-PCR, reverse transcription polymerase chain reaction
Data in parentheses are 95% confidence intervals.

In samples from symptomatic patients, 32 of 36 reference RT-PCR–positive samples were also positive by antigen testing (Table 3a). The sensitivity, specificity, PPV, and NPV were 88.9% (95% CI: 73.9–96.9%), 100% (95% CI: 99.3–100%), 100% (95% CI: 84.2–100%), and 99.5% (95% CI: 98.8–99.9%), respectively.

**Table 3a.** **Sensitivity and specificity of QuickChaser**^**®**^ **Auto SARS-CoV-2 among symptomatic patients**

| Symptomatic subjects |  | Real-time RT-PCR |  |
| --- | --- | --- | --- |
|  |  | Positive | Negative |
| QuickChaser<sup>®</sup> Auto SARS-CoV-2 | Positive | 32 | 0 |
|  | Negative | 4 | 825 |
| Sensitivity (%) |  | 88.9(73.9–96.9) |  |
| Specificity (%) |  | 100 (99.3–100) |  |
| Positive predictive value (%) |  | 100 (84.2–100) |  |
| Negative predictive value (%) |  | 99.5 (98.8–99.9) |  |

RT-PCR, reverse transcription polymerase chain reaction
Data in parentheses are 95% confidence intervals.

In samples from asymptomatic individuals, 30 of 47 reference RT-PCR–positive samples were positive by antigen testing (Table 3b). The sensitivity, specificity, PPV, and NPV were 63.8% (95% CI: 48.5–77.3%), 99.6% (95% CI: 98.5–100%), 93.8% (95% CI: 79.2–99.2%), and 96.7% (95% CI: 94.7–98.0%), respectively.

**Table 3b.** **Sensitivity and specificity of QuickChaser**^**®**^ **Auto SARS-CoV-2 among asymptomatic patients**

| Asymptomatic subjects |  | Real-time RT-PCR |  |
| --- | --- | --- | --- |
|  |  | Positive | Negative |
| QuickChaser<sup>®</sup> Auto SARS-CoV-2 | Positive | 30 | 2 |
|  | Negative | 17 | 491 |
| Sensitivity (%) |  | 63.8 (48.5–77.3) |  |
| Specificity (%) |  | 99.6 (98.5–100) |  |
| Positive predictive value (%) |  | 93.8 (79.2–99.2) |  |
| Negative predictive value (%) |  | 96.7 (94.7–98.0) |  |

RT-PCR, reverse transcription polymerase chain reaction
Data in parentheses are 95% confidence intervals.

The sensitivities of the antigen test stratified by Ct value are shown in Table 4.

**Table 4.** **QuickChaser^®^ Auto SARS-CoV-2 sensitivities stratified by the Ct values**

| Ct value (N2 gene) | Antigen test |  | Sensitivity |  |
| --- | --- | --- | --- | --- |
|  | Positive<br>(n = 62) | Negative<br>(n = 21) |  |  |
| < 20 | 16 | 0 | 100.0% | (71.3–100%) |
| 20-24 | 28 | 1 | 96.6% | (82.2–99.9%) |
| 25-29 | 13 | 0 | 100.0% | (66.1–100%) |
| ≥ 30 | 5 | 20 | 20.0% | (6.8–20.7%) |

Data in parentheses are 95% confidence intervals.
Ct, cycle threshold

### Detailed data of samples with discrepant results between QuickChaser^®^ Auto SARS-CoV-2 and reference RT-PCR assay

Of the 23 discrepant samples, two were positive by the antigen test and negative by reference RT-PCR (false positive). Of the 21 samples that were negative by the antigen test and positive by reference RT-PCR (false negative), 20 had Ct values >30, and one had a Ct value of 20. For a patient with a false-negative result despite a Ct value of 20, we retested the preserved UTM™ sample with QuickChaser^®^ Auto SARS-CoV-2. The antigen test provided a positive result for UTM™ samples that were diluted approximately 40 fold.

## Discussion

In this prospective study, Quick Chaser^®^ Auto SARS-CoV-2 using nasopharyngeal specimens demonstrated a sensitivity of 74.7% and a specificity of 99.8%. In patients with Ct values <30, the sensitivity was almost identical to RT-PCR. Furthermore, Quick Chaser^®^ Auto SARS-CoV-2 had a lower LOD than other antigen tests currently approved in Japan.

Several antigen tests have been developed, with generally high specificity and variable sensitivity [5]. While Quick Chaser^®^ Auto SARS-CoV-2 had the lowest LOD in this study, QuickNavi [8] and Panbio [10] had comparable sensitivities. The diagnostic performance of antigen tests may differ between experimental (UTM™) and clinical samples [11]. Therefore, direct comparison using clinical samples should be conducted to evaluate the real-life performance of each test.

Viral load influences overall sensitivity, as shown by the fact that the sensitivity of antigen tests generally plummets in samples with Ct >30 [8,12]. Samples with Ct >30 comprised 30.1% (25/83) of our study population, which may have decreased the overall sensitivity of Quick Chaser^®^ Auto SARS-CoV-2. Another factor that may influence sensitivity is the swab type used. Quick Chaser^®^ Auto SARS-CoV-2 includes sponge swabs, although flocked-type swabs can collect samples more efficiently [13]. Despite the aforementioned challenges, this antigen test successfully detected SARS-CoV-2 in all but one of the samples with Ct <30. The remaining case was a false negative, and considering that re-examination with the UTM™ sample showed a positive result, the original finding may have been caused by a low viral concentration due to a flawed sample collection technique. The good performance of this test indicates that it can accurately identify contagious patients, given that those with Ct values <30 are considered to be highly infectious [14].

We observed false positives in only two samples, and the specificity of Quick Chaser^®^ Auto SARS-CoV-2 was over 99%. False positives should be avoided as they lead to unnecessary further testing or quarantine measures [15]; thus, the specificity is recommended to be over 97% [5]. Positive results should be cautiously interpreted, especially when the prevalence of SARS-CoV-2 or possibility of infection is low.

To maximize its sensitivity, Quick Chaser^®^ Auto SARS-CoV-2 uses two methods: silver amplification and digital interpretation of the results. Similar to many antigen tests [5], Quick Chaser^®^ Auto SARS-CoV-2 implements sandwich methods using labeled antibodies and capture antibodies. Antibodies labeled with gold colloid attach to specific antigens in a sample. The labeled antigens are then sandwiched by capture antibodies, which indicate the positive bands. The silver amplification method generates large silver particles using the gold colloid as a catalyst, and thus enhances the visibility of the labeled antibody complex [7]. A previous study showed that among antigen tests for the influenza virus, those that used this method had higher sensitivity than those that did not (type A, 91.2% vs. 86.8%, respectively; type B, 94.4% vs. 81.2%, respectively), without compromising specificity [16]. Similarly, digital scans of the test reagents can improve the accuracy of antigen detection. Digital scans also increase the objectivity of test result interpretation by removing the necessity for visual inspection. A systematic review suggested that digital immunoassays increased sensitivities by 25.5% and 23.5% for influenza A and B, respectively [17]. Although digital immunoassays require special equipment, the additional cost is much cheaper than that of NAATs, and is compensated for by the increase in sensitivity [17].

There are several limitations regarding this study. First, reference real-time RT-PCR used frozen samples. While samples were stored at -80 °C, their viral load may have decreased during storage process. Second, we did not investigate whether mutations in SARS-COV-2 affected the diagnostic performance. Third, we did not evaluate saliva or anterior nasal cavity samples. Saliva collection and anterior nasal swabs cause less pain and coughing than nasopharyngeal swabs [18]. Future studies should compare the diagnostic performance of samples obtained using each of these methods.

In conclusion, Quick Chaser^®^ Auto SARS-CoV-2 showed satisfactory diagnostic performance of symptomatic patients. The sensitivity was especially high in samples of Ct <30, indicating that the test can accurately detect highly infectious patients.

## Data Availability

The data that support the findings of this study are available from the corresponding author, Y.K., upon reasonable request.

## Acknowledgments

We thank the staff in the Department of Clinical Laboratory of Tsukuba Medical Center Hospital for their intensive support of this study. We thank all of the medical institutions for providing their patients’ clinical information.

